# The German COVID-19 Survey on Mental Health: Primary Results

**DOI:** 10.1101/2020.05.06.20090340

**Authors:** Stefanie Jung, Jonas Kneer, Tillmann H.C. Kruger

## Abstract

First cases of COVID-19 were reported in Wuhan, China in early December 2019. Preliminary data from China indicated that the pandemic and its associated lockdown measures may have a substantial impact on mental health and well-being, with evidence of increased levels of psychological distress, anxiety, depressive symptoms and insomnia.^1,2^ In March 2020, the German government agreed upon a substantial catalogue of measures including contact bans that came into effect on 22 March. Such measures are unprecedented for the majority of people and may affect their lives tremendously. Thus, the current survey was immediately developed to systematically assess mental health in response to these measures.

**Methods:** The survey was approved by the local ethics committee at Hannover Medical School, Germany and included web-based self-report measures as outlined below. First wave data were taken during the height of lockdown measures in Germany from 1 April to 15 April 2020.

**Results:** *Demographics:* A total of 3,545 volunteers took part in this cross-sectional survey. Mean age was 40.36 years (SD = 11.70; 83.1% female, 15.2% male), mean educational years 15.87 (SD = 4.19), 9.9% were unemployed and 23.9% reported living alone. Acute or chronic disease was reported by 36.7% (physical) and 24.7% (mental) of subjects.

*Distress, Anxiety and Depression:* Psychosocial distress as measured with the PHQ stress module (items 12a-12j of PHQ-D) was at M = 6.36 (SD = 0.89), implying mild psychosocial distress (range 5-9). Depression and anxiety as assessed by PHQ-4 was at M = 3.80 (SD = 3.03) and significantly higher than in a reference sample (t(6008) = 32.78, p = 0.00).^3^ The mean well-being score (WHO-5) was 50.7 (SD = 23.8) (range 0-100), with normal individuals having a mean score of 75 and subjects with major depression 37.5.^4^ The majority of subjects (60%) indicated very good or fair, 26.9% poor or very poor subjective coping with the pandemic and corresponding measures. Calculation of gender differences revealed higher scores for depression and anxiety (t(3459) = 4.93, p = 0.00) and poorer coping in women (U = 678156, p = 0.00).

*Sleep, irritability & violence:* Using comparative questions on a 5-point Likert scale 45.3% of participants reported worsened sleep compared to pre-pandemic times. Of all participants 50.9% reported being more easily irritated (compared to 12.2% feeling less easily irritated) and 29% reported experiencing more anger and aggression (compared to 12.8% experiencing less). Of these 65.5% directed their anger and aggression at others, while 32.6% directed it at themselves. Most importantly, 5% of all participants reported experiencing interpersonal violence (IPV) on a verbal (98.4%), physical (41.9%) or sexual (30.2%) level. In case of verbal violence, 77.3% reported experiencing more verbal violence lately (compared to 3.4% experiencing less). Regarding physical violence, 19.5% reported experiencing increased levels (compared to 2.8% experiencing less) and in case of sexual violence more people reported experiencing increased sexual violence lately (11.1%) compared to 1.7% that experienced less.

**Discussion:** This is one of the first and largest surveys on mental health during COVID pandemic in a European society. Although the cohort reflects a relatively well educated and financially secure sample, there is evidence of substantial mental burden with increased levels of stress, anxiety, depressive symptoms, sleep disturbance and irritability. Most importantly and also most concerning is the finding of a one-month prevalence of 5% IPV, which is already close to one-year prevalence rates^5^ and for which there were indices that this has currently increased. We think it is of vital importance to continuously monitor the mental health of the general public during this pandemic and its aftermath and to carefully screen for IPV and its risk factors such as stress, sleep problems and anger.^6^

## Methods

The survey was approved by the local ethics committee at Hannover Medical School, Germany and included web-based self-report measures as outlined below. First wave data were taken during the height of lockdown measures in Germany from 1 April to 15 April 2020.

## Results

### Demographics

A total of 3,545 volunteers took part in this cross-sectional survey. Mean age was 40.36 years (SD = 11.70; 83.1% female, 15.2% male), mean educational years 15.87 (SD = 4.19), 9.9% were unemployed and 23.9% reported living alone. Acute or chronic disease was reported by 36.7% (physical) and 24.7% (mental) of subjects.

### Distress, Anxiety and Depression

Psychosocial distress as measured with the PHQ stress module (items 12a-12j of PHQ-D) was at M = 6.36 (SD = 0.89), implying mild psychosocial distress (range 5-9). Depression and anxiety as assessed by PHQ-4 was at M = 3.80 (SD = 3.03) and significantly higher than in a reference sample (t(6008) = 32.78, p = 0.00).^3^ The mean well-being score (WHO-5) was 50.7 (SD = 23.8) (range 0-100), with normal individuals having a mean score of 75 and subjects with major depression 37.5.^4^ The majority of subjects (60%) indicated very good or fair, 26.9% poor or very poor subjective coping with the pandemic and corresponding measures. Calculation of gender differences revealed higher scores for depression and anxiety (t(3459) = 4.93, p = 0.00) and poorer coping in women (U = 678156, p = 0.00).

### Sleep, irritability & violence

Using comparative questions on a 5-point Likert scale 45.3% of participants reported worsened sleep compared to pre-pandemic times. Of all participants 50.9% reported being more easily irritated (compared to 12.2% feeling less easily irritated) and 29% reported experiencing more anger and aggression (compared to 12.8% experiencing less). Of these 65.5% directed their anger and aggression at others, while 32.6% directed it at themselves. Most importantly, 5% of all participants reported experiencing interpersonal violence (IPV) on a verbal (98.4%), physical (41.9%) or sexual (30.2%) level. In case of verbal violence, 77.3% reported experiencing more verbal violence lately (compared to 3.4% experiencing less). Regarding physical violence, 19.5% reported experiencing increased levels (compared to 2.8% experiencing less) and in case of sexual violence more people reported experiencing increased sexual violence lately (11.1%) compared to 1.7% that experienced less.

## Discussion

This is one of the first and largest surveys on mental health during COVID pandemic in a European society. Although the cohort reflects a relatively well educated and financially secure sample, there is evidence of substantial mental burden with increased levels of stress, anxiety, depressive symptoms, sleep disturbance and irritability. Most importantly and also most concerning is the finding of a one-month prevalence of 5% IPV, which is already close to one-year prevalence rates^5^ and for which there were indices that this has currently increased. We think it is of vital importance to continuously monitor the mental health of the general public during this pandemic and its aftermath and to carefully screen for IPV and its risk factors such as stress, sleep problems and anger.^6^

## Data Availability

All published data are available.

## Notes

### Competing Interest Statement

The authors have declared no competing interest.

### Funding Statement

No external funding was received.

